# A collaborative network trial to evaluate the effectiveness of implementation strategies to maximise adoption of a school-based healthy lunchbox program: A study protocol

**DOI:** 10.1101/2023.11.19.23298746

**Authors:** Courtney Barnes, Jannah Jones, Luke Wolfenden, Katie Robertson, Anna Lene Seidler, Jennifer Norman, Pip Budgen, Megan Mattingly, Carla Piliskic, Lisa Moorhouse, Jennifer Mozina, Jennifer Plaskett, Sarah McDermott, Sara Darney, Cecilia Vuong, Nina Douglass, Kara McDonnell, Rachel Sutherland

## Abstract

**Background:** Schools provide universal access to children over five years of age, representing a key opportunity for nutrition interventions to prevent the development of chronic disease. However, an important impediment to the large-scale adoption of evidence-based school nutrition interventions is the lack of evidence on effective strategies to implement them. This paper describes the protocol for a “Collaborative Network Trial” to support the simultaneous testing of different strategies undertaken by New South Wales Local Health Districts to facilitate the adoption of an effective school-based healthy lunchbox program (‘SWAP IT’). The primary objective of this study is to assess the effectiveness of different implementation strategies to increase school adoption of the SWAP across New South Wales Local Health Districts.

**Methods:** Within a Master Protocol framework, a collaborative network trial will be undertaken. Independent randomised controlled trials to test implementation strategies to increase school adoption of SWAP IT within primary schools in 10 different New South Wales Local Health Districts will occur. Schools within each Local Health District will be randomly allocated to either the intervention or control condition. Schools allocated to the intervention group will receive a combination of implementation strategies developed by each of the Local Health Districts independently, based on their existing capacities and local contexts. Across the 10 participating Local Health Districts, six broad strategies were developed and combinations of these strategies will be executed over a 6 month period. In six districts an active comparison group (containing one or more implementation strategies) was selected. The primary outcome of the trial will be adoption of SWAP IT, assessed via electronic registration records captured automatically following online school registration to the program. The primary trial outcome, between-group differences at 6 month follow-up, will be assessed using logistic regression analyses for each trial. Individual participant data component network meta-analysis, under a Bayesian framework, will be used to explore strategy-covariate interactions; to model additive main effects (separate effects for each component of an implementation strategy); two way interactions (synergistic/antagonistic effects of components), and full interactions.

**Discussion:** The study will provide rigorous evidence of the effects of a variety of implementation strategies, employed in different contexts, on the adoption of a school-based healthy lunchbox program at scale. Importantly, it will also provide evidence as to whether health service-centred, collaborative research models can rapidly generate new knowledge and yield health service improvements.

## INTRODUCTION

Dietary risk factors are a leading cause of preventable death and disability. (1) Reducing dietary risks is recommended to improve child health and mitigate future burdens of chronic disease.(2) In Australia, for example, 96% of children do not consume sufficient serves of vegetables, whilst discretionary foods (i.e. foods high in added sugar, saturated fat and sodium) account for over one-third of children’s daily energy intake.(3) Schools provide universal access to children aged over 5 years, and are a setting recommended for nutrition interventions in chronic disease prevention internationally.(4–6) In countries such as Australia, food brought to school (from home) packed in school ‘lunchboxes’ are used daily by 90% of students,(7) and contribute up to 30-50% of a child’s daily energy intake.(7) As approximately 40% of foods in lunchboxes are discretionary(8) improving the packing of healthy foods for child consumption at school provides a considerable opportunity for chronic disease prevention.

Systematic reviews suggest that school-based healthy lunchbox interventions can improve student nutritional intake.(9) In Australia, a series of randomised controlled trials of a healthy lunchbox program, known as ‘SWAP IT’ were recently conducted in 34 primary schools with 4600 children.(10, 11) The program supports parents and carers to make simple ‘swaps’ aligned to dietary guidelines,(12) replacing discretionary food and beverage items with comparable core (nutrient dense) items. It is comprised of three broad program components: i.) school food (lunchbox) guidelines; ii.) messages and hard copy resources to parents and carers; and iii.) curricula resources for teachers. Across these randomised trials, the program was found to significantly improve child diet quality, energy intake and weight status, and was acceptable to both parents and teachers.(10, 11) A subsequent comparative effectiveness randomised trial found no difference in effectiveness between the messages and parent booklets combined, compared with those two components plus school-based curriculum and policy resources on student dietary outcomes.

Given the reported benefits of SWAP IT on child health,(10, 11) broad implementation in schools has the potential to make a significant contribution to improving public health nutrition. An important impediment to the large scale adoption of effective school nutrition initiatives, however, is a lack of published evidence of effective strategies to implement them.(13) A recent Cochrane review of implementation strategies for school-based health promotion programs identified few randomised controlled trials of strategies to implement policies and practices promoting healthy eating, particularly ‘at scale’ (defined by the authors as 50 or more schools).(13) Furthermore, strategies identified as effective in improving implementation in one jurisdiction (e.g. Local Health District), may not be effective, appropriate or feasible for application in another. Similarly, differing capacities (e.g. resources or infrastructure) of agencies responsible for undertaking or supporting program implementation may mean an effective implementation strategy in one jurisdiction may not be feasible to execute in another. Such issues must be addressed if effective interventions are to be adopted at a population level (at scale).

As in clinical services, systematic reviews and best practice guidelines identify evidence-based programs and practices that can be employed in community settings to reduce child dietary risks. As such, within devolved health systems such as Australia, different health services will often seek to address the same disease risk or health condition, using the same intervention (e.g. guideline concordant care) at the same time.(14) These services, however, operate in different contexts, with different capacities and resource constraints. As a result, there is often natural heterogeneity in the strategies that health services employ to support the implementation of programs in schools and other clinical and community settings to improve dietary (and other) outcomes. This convergence of objective (to implement a similar intervention), but heterogeneity in context and strategies used to implement school-based programs, presents an attractive opportunity to learn about the types of implementation strategies that may be effective in different contexts. Specifically, the coordinated evaluation of implementation efforts across a network of health services, and the establishment of processes to share and learn from the findings, may provide a mechanism for rapid evidence generation, and health system improvement ‘at scale’. Such collaborative and data-driven models of working are also consistent with recommendations for the development of ‘learning health system’ approaches to healthcare improvement.(15)

Broadly, Master Protocols represent an approach that could be used to facilitate coordinated and collaborative research, learning and improvement.(16) Typically used to test pharmacological interventions, they refer to designs employing coordinated approaches to assess the effects of interventions within a unifying overall trial structure.(16) This infrastructure, including a centralised trial protocol and governance, facilitates the standardisation of study processes and procedures, including recruitment, evaluation and data collection, analysis, and reporting.(17) They allow for the examination of multiple hypotheses,(18) such as the effects of a variety of implementation or scale-up strategies on school adoption of health promotion programs, or differences in effectiveness for different population groups. Following demonstration of the effectiveness and acceptability of the SWAP IT program,(10, 11) three Local Health Districts (LHDs) from across New South Wales (NSW), Australia, expressed interest in supporting the implementation of this program in their LHD. In this context, and drawing on research design principles of Master Protocols and prospective meta-analysis methodology,(19) a pilot collaboration was formed that networked three LHDs and the University of Newcastle (National Centre of Implementation Science)(20) to undertake a harmonised evaluation of strategies used within each LHD to support the adoption of the SWAP IT program, and to share learning from these evaluations across participating LHDs.(21) The collaboration was supported by shared implementation strategy development processes, governance structures, centralised data collection infrastructure, and a community of practice.(21) Whilst collaboration across and flexibility within LHDs for the implementation of various health promotion programs has occurred routinely amongst NSW LHDs over time, a formal evaluation of such a collaborative approach had not been undertaken. The pilot found the collaborative model was highly acceptable to all parties,(21) and strategies employed yielded significant, but contextually dependent improvements in program adoption.

Based on these encouraging findings, the collaborative approach is now being employed across 10 of the 15 LHDs (67%) in NSW. This paper describes the protocol for what we term a “Collaborative Network Trial” to support the simultaneous testing of different implementation strategies undertaken by 10 LHDs in NSW, Australia to facilitate the adoption of the SWAP IT program at scale.

## OBJECTIVES

As such, the primary objective of this study is to assess, using individual level participant (in this case ‘school’) data (IPD), the effectiveness of different implementation strategies employed by 10 NSW LHDs to increase school adoption of the SWAP IT program.

Secondary objectives of the study are to: (1) explore the effects of different implementation strategy components and contextual factors on the school-level adoption of SWAP IT using pooled individual level data across all trials; (2) assess the acceptability of the implementation strategies to school principals; and (3) assess the sustainability of SWAP IT within schools that adopted the program at 18-months.

## METHODS

### Context

LHDs are NSW Government funded health services responsible for providing or supporting the provision of health promotion services to address the leading risk factors for chronic disease in their community. The NSW Ministry of Health provides funding to LHDs to support the implementation of state-wide health promotion programs.(22) All NSW LHDs have received funding to facilitate the implementation of healthy eating and physical activity policies and practices in NSW primary schools for over a decade as part of the NSW Healthy Children’s Initiative.(22) However, although healthy lunchboxes have historically been a focus for health promotion activities in some LHDs and non-government organisations (e.g. Cancer Council NSW), the funding provided by NSW Ministry of Health did not explicitly focus on a formal school-based program to support the packing of healthy lunchboxes. In addition, whilst a core component of health promotion practice, Health Promotion Unit capability to undertake research and evaluation of health promotion activity has been found to vary across LHDs.(23)

### Ethics and trial registration

The research will be conducted and reported in accordance with the requirements of the Consolidated Standards of Reporting Trials (CONSORT) Statement.(24) Ethics approval has been obtained via the following Human Research Ethics Committees: Hunter New England (2019/ETH12353); University of Newcastle (09/07/26/4.04); NSW of Department of Education (2018247); and the Maitland-Newcastle, Sydney, Wollongong, Bathurst, Parramatta, Wagga Wagga and Canberra-Goulburn Catholic Dioceses. This trial is also registered prospectively with the Australian New Zealand Clinical Trials Registry (ACTRN12623000558628).LJThe protocol is reported according to the Standard Protocol Items: Recommendations for Interventional Trials (SPIRIT) (Supplementary File 1).(25)

### Study design and setting

Within a Master Protocol framework,(16) we will undertake a Collaborative Network Trial. Specifically, independent randomised controlled trials to test strategies to implement or improve health care occurring at different sites (LHDs) will be undertaken by the Health Promotion Units at each LHD. The key trial methods, measures and data collection processes will be harmonised with agreement across sites to provide individual school-level data for planned pooled analyses as part of a collaborative, following a prospective meta-analysis framework.(19) The design allows for heterogeneity or natural variation in the implementation strategies being tested and the contexts (i.e. sites) they are tested in.(16) The study builds on a pilot network trial to implement the scale-up of SWAP IT program in three LHDs.(13)

### Sample and participants

The study will be conducted with primary and combined schools located across 10 LHDs in NSW, Australia. The state of NSW has approximately 2100 primary and combined schools, and is socioeconomically and geographically diverse.(26) Department of Education (DoE), Catholic Schools NSW and Association of Independent Schools of NSW primary and combined schools located within the LHDs of Murrumbidgee, Hunter New England, Sydney, Western Sydney, South Western Sydney, South Eastern Sydney, Northern Sydney, Western NSW, Nepean Blue Mountains, and Illawarra Shoalhaven will be included in the study.

Primary and combined schools located within the participating LHDs who cater for at least one primary school year and have not implemented the SWAP IT program will be eligible to participate. Only schools that do not use the Audiri parent communication app will be eligible, as these schools are participating in another trial being conducted concurrently by the research team (ACTRN12623000145606). Schools catering exclusively for children requiring specialist care, for example schools catering for students with severe disabilities, will be excluded. Schools with secondary students only will also be ineligible to participate.

All eligible schools will be included in the study as part of usual service delivery provided by LHD health promotion staff to support schools to implement a range of health promotion programs. Eligible schools will be invited to participate in the secondary data collection component of the study, specifically the follow-up survey conducted with school principals (described below). Schools will be recruited for the follow-up data collection via an invitation email containing a link to an online survey and a study information statement outlining the purpose of the research and their involvement. Schools that are yet to complete the survey will receive up to three reminder prompts via telephone or email by the research team to encourage completion.

### Randomisation and blinding

Prior to the delivery of the first scale-up strategy, schools within each LHD will be randomly allocated to either the intervention or control condition using a computerised random number function in a 1:1 (intervention: control) ratio. Randomisation will be stratified by school size and social socio-economic location, as determined by Socio-Economic Indexes for Areas categorisation using school postcodes,(27) given the socio-economic association with implementation of school nutrition programs.(28) Randomisation will be completed by a statistician not otherwise involved in the trial. Due to the nature of the intervention, participants will not be blinded to group allocation. However, research staff assessing the outcomes at follow-up will be blinded.

### Implementation strategies

Implementation strategies were developed for each of the LHDs independently, based on their existing capacities and local contexts. Implementation strategies for each participating LHD (‘site’) were co-designed by LHD health promotion staff and other stakeholders, with support provided by National Centre of Implementation Science (NCOIS) implementation scientists and SWAP IT developers from the University of Newcastle. The development process included: i.) planning workshops facilitated by University staff that drew on tacit knowledge and experience of health promotion staff who had considerable experience working with schools; ii) evidence regarding barriers to school adoption and implementation of SWAP IT collected by the research team as part of previous SWAP IT trials, iii) data from systematic reviews and pilot trials regarding the effectiveness of strategies to facilitate adoption.(29) During the workshops, theoretical framework tools were used to facilitate the selection of strategies to address barriers that were aligned to individual LHD capacity and contexts.(30–32) Processes may have also been undertaken by LHDs to identify strategies to support access and engagement of priority populations within their region to ensure school adoption and implementation of SWAP IT does not further exacerbate health inequities. This may have included consultation and engagement processes with Aboriginal, or Culturally and Linguistically Diverse individuals, groups or stakeholders.

Across the 10 participating LHDs, six broad implementation strategies emerged and are described below. The combination of these six strategies employed by each LHD will differ and is described in Table 1. For all LHDs, these strategies will be executed over a period of six months. A timeline for the delivery of the implementation strategies is provided in Table 2.

**1. Sector support and endorsement:** Policy makers from Health will target principals to communicate, support and endorse the program and its outcomes, its alignment to sector policies and recommend its adoption. This endorsement will occur via a maximum of two targeted letters or emails developed by the research team, approved and endorsed by local and state-level Health partners. The letters or emails will also contain a link to resources and the enrolment website. As an additional strategy, some LHDs (outlined in Table 1) will use their existing connections to obtain endorsement for the program from local educational and wellbeing liaisons within the NSW Department of Education. This endorsement will be promoted to schools via an email distributed by the liaisons directly to schools receiving this strategy.
**2. Local facilitation:** Health promotion staff from LHDs have developed strong and trusted local relationships with schools for over a decade and represent credible sources of local nutrition expertise. LHD health promotion staff will use up to two of their existing planned school contacts, conducted via telephone call or face-to-face meeting, to assess interest in the SWAP IT program, address any school-specific barriers to adoption, and facilitate goal setting and action planning. Scripts developed by the research team to guide the local facilitation will incorporate motivational interviewing techniques to be employed by health promotion staff to address school barriers to program adoption.
**3. Develop and distribute educational materials**: Targeted at principals to address perceived barriers to adoption, the strategy will initially aim to create tension for change (e.g. via outlining parent and carer interest and expectations); and then communicate the attractive program attributes (e.g. simplicity, no-cost). This communication will consist of up to two contacts, including a printed information pack (consisting of a flyer, SWAP IT pen and example parent booklet) at the commencement of the intervention period followed by an email to promote the program. As an additional strategy, one LHD will offer printed parent booklets promoting the SWAP IT program to all parents and carers with children commencing the following school year within their school kindergarten orientation packs along with a flyer encouraging the school principal or wellbeing coordinator to adopt the program.
**4. Local opinion leaders**: Promotional materials, including one printed information pack (consisting of a flyer and example SWAP IT parent booklet) and one email, will be delivered to other leaders that may be influential in a schools decision to adopt health promotion programs, specifically the school administration manager and parent committee. The aim of these materials is to promote the SWAP IT program and encourage school adoption.
**5. Audit and feedback:** Data and feedback on school adoption of SWAP IT will be automatically captured through electronic registration records and be provided to schools via other implementation strategies, including educational materials, local facilitation and local opinion leaders. For example, educational materials provided to principals, school administration managers and parent committees will include information on the number of schools that have registered for SWAP IT, a link to view an online list of schools have already adopted the program (to create tension for change and social norms) and provide instruction on how the school can also register for the program.
**6. Educational meeting:** Health promotion staff from LHDs will conduct one webinar with schools within their LHD to assess interest in the SWAP IT program and address any barriers to adoption. Webinar content will be developed by the research team in collaboration with health promotion staff.

**Table 1.**
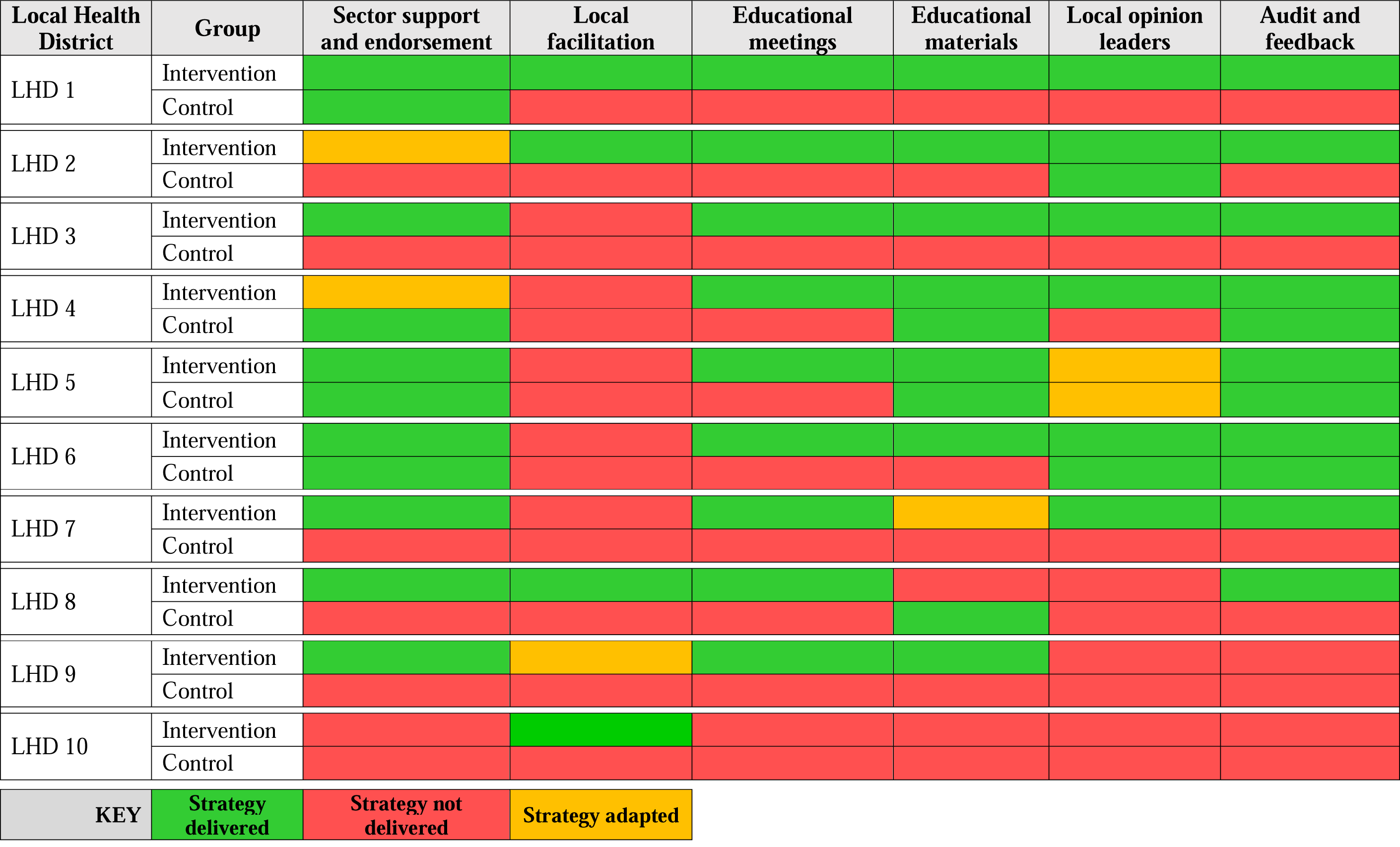
Implementation strategies delivered by each Local Health District.

**Table 2.**
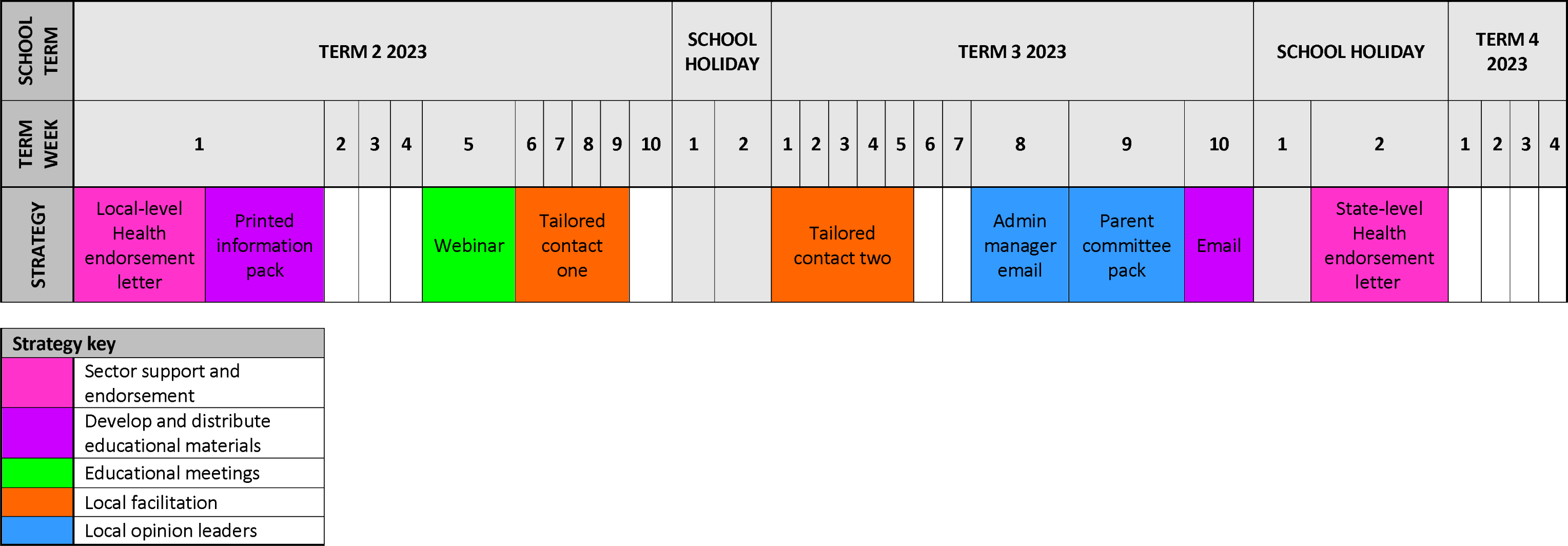
Timeline for the delivery of the implementation strategies.

### Control group and contamination

Registration for the SWAP IT program is publicly available and freely accessible for all schools, including schools allocated to the control group. The implementation strategies to be delivered to the control group across LHDs is described in Table 1. For most schools allocated to the control group, the comparison will be ‘no implementation support’ or a singular strategy. Execution of the implementation strategies will be monitored centrally by the research team in consultation with health promotion staff from each LHD to minimise risk of contamination. Nonetheless, school exposure to the implementation strategies will be assessed at follow-up via an online or telephone survey with school principals (described below).

### Study outcomes and data collection

Trial outcomes were discussed and agreed upon by participating LHDs. Data collection for all trial outcomes were harmonised across all LHDs and will be collected centrally by the research team at the University of Newcastle. The centralisation of data collection represented an efficient means of collecting and managing data for all participating LHDs. All demographic, operational and trial outcome measures are harmonised (i.e. identical item, measure and data collection method) to facilitate comparability and analysis. Each participating LHD will retain access to their trial dataset.

### Primary outcome

<u>Adoption of the SWAP IT program</u>, defined as the number of schools who register for the lunchbox nutrition program (SWAP IT), will be assessed within schools allocated to the intervention and control group via electronic registration records captured automatically following school registration to SWAP IT. As part of the registration process, schools provide consent for the de-identified registration data to be used for research and evaluation purposes. This outcome will be assessed at baseline and approximately 9 months after baseline data collection.

### Secondary outcomes

<u>Acceptability of implementation strategies,</u> defined as the perception amongst principals that the implementation strategies are agreeable, palatable or satisfactory,(33) will be assessed in a telephone or online survey with school principals at 9-month follow-up. School principals will be asked if they recall receiving each of the implementation strategies during the intervention period. For strategies the participants recall receiving, they will be asked to rate how acceptable they found the strategy on a 5-point Likert scale (1=not acceptable; 5=very acceptable). Principals from 243 Catholic and Independent primary schools located across five LHDs (LHD 1; LHD 5; LHD 7; LHD 8; LHD 9) will be invited to participate in the survey. These LHDs have been selected as they are employing diverse combinations of the implementation strategies (Table 1). Including schools from these LHDs in the survey will ensure the acceptability of all employed strategies (across the 10 LHDs) will be assessed without surveying all participating schools.

<u>Implementation of the SWAP IT program</u>, defined as the extent to which the SWAP IT program components were delivered by the school to parents, will be assessed in the telephone or online survey with school principals at 9-months follow-up. Schools will be asked to report if they implemented the SWAP IT program at their school, and what program components were implemented (i.e. parent messages; school lunchbox guidelines; curriculum resources; parent and carer resources).

Sustainability of the SWAP IT program, defined as continued school use of the lunchbox nutrition program (SWAP IT) at 18 months after baseline data collection, will be assessed via electronic registration records captured automatically following school registration to SWAP IT.

<u>School characteristics,</u> including postcode, total student enrolments, geographic location (urban, regional, rural and remote), proportion of Aboriginal student enrolments, and proportion of students that speak a language other than English at home, were obtained from a publically accessible Australian Curriculum, Assessment and Reporting Authority (ACARA) database.(34)

### Sample size and data analysis

We are anticipating a sample of at least 30 schools per group (and an average of 60 per group) in trials of each of the 10 participating LHDs. Descriptive statistics, including proportions, means and standard deviations, will be used to describe school characteristics, adoption, implementation and sustainability of SWAP IT, as well as the acceptability of the implementation strategies.

Analyses of trial outcomes will be undertaken under an intention to treat framework separately for each trial. For assessment of school level program adoption, the primary trial outcome, between-group differences, will be assessed using logistic regression. The model will include a term for treatment group (intervention vs control) and pre-specified covariates prognostic of the outcome. Little, if any, missing primary outcome data is anticipated at follow-up, as program adoption is recorded automatically for all participating schools.

Nonetheless, we will employ multiple imputation for any missing data in the event that schools withdraw from the study and request that their data are not used. All statistical tests will be 2 tailed with alpha of 0.05. Assuming adoption of the program by 10% in the comparison group, a sample size of approximately 30 schools per group will be sufficient to detect an absolute difference between groups of 30%, with 80% power and an alpha of 0.05.

We will employ component IPD component network meta-analysis to compare and rank the effects from all the tested strategies on the primary trial outcome.(35) For this analysis we will also include the three randomised controlled trials from the pilot,(21) expanding the network and providing pooled individual level data from 13 randomised controlled trials. We will explore combining ‘educational meetings and educational materials’ into a single component for analysis given their shared underlying behavioural targets. We will adjust for prognostic factors and exploration of strategy—covariate interactions to identify if and to what extent effects vary by participant, population or other contextual factors (effect modifier).(35) We will also employ component network meta-analyses to model additive main effects (separate effects for each element or component of an implementation strategy); two way interactions (synergistic/antagonistic effects of components), and full interactions (different effects from each combination of components). The analyses will be performed under a Bayesian framework. There are no established methods for sample size calculations for component network meta-analysis.

### Trial governance

The trial will be overseen by a Steering Group, comprised of representatives from each LHD, including: Aboriginal Health Promotion Managers; program developers, implementation scientists, trialists and research dietitians from the University of Newcastle. Roles and responsibilities will be documented in a Terms of Reference for the Group. LHDs will be responsible for the selection of implementation strategies for their jurisdiction, and execution of some of the strategies to schools. The University of Newcastle will be responsible for facilitating trial workshops, ethics, data collection, monitoring and quality assurance, data management and analysis. A Community of Practice, established in the pilot,(21) will also be employed to support the interpretation of trial results and pooled analyses, exchange tacit knowledge and experience and identify opportunities for improvement.

## DISCUSSION

This protocol provides a comprehensive description of a novel research design to help generate evidence that can better inform approaches to support the adoption and implementation of health promotion interventions at scale. The study will provide rigorous evidence of the effects of a variety of implementation strategies, employed in different contexts on the adoption of the SWAP IT school lunchbox program. Importantly, it will also provide published evidence whether health service-centred, collaborative research models can rapidly generate new knowledge and yield health service improvements.

## Data Availability

All data produced in the present study are available upon reasonable request to the authors

## ACKNOWLEDGEMENTS

We authors would like to thank the health promotion staff from all participating Local Health Districts for their contributions to the project. We would also like to thank Sol Libesman and Christophe Lecathelinais for statistical support. Finally, we would like to acknowledge the National Centre of Implementation Science, a National Health and Medical Research Centre for Research Excellence (APP1153479), for the support provided to the study.

## FUNDING

Courtney Barnes receives salary support from a NSW Ministry of Health PRSP Research Fellowship. Luke Wolfenden is supported by an NHMRC Investigator Grant (APP11960419) and NSW Cardiovascular Research Capacity Program (H20/28248). Rachel Sutherland is supported by a Medical Research Future Fund Fellowship (APP1150661) and a Hunter New England Clinical Research Fellowship. Jennifer Norman receives salary support from the NSW Ministry of Health PRSP funding awarded to Early Start at the University of Wollongong. Jannah Jones is supported by a Hunter New England Clinical Research Fellowship. Anna Lene Seidler is supported by an NHMRC Investigator Grant (APP2009432). The contents of this manuscript are the responsibility of authors and do not reflect the views of NSW Ministry of Health or NHMRC.

## COMPETING INTERESTS

All authors declare that they have no competing interests.

## ETHICS DECLARATIONS

### Ethics approval and consent to participate

Ethics approval has been obtained via the following Human Research Ethics Committees: Hunter New England (2019/ETH12353); University of Newcastle (09/07/26/4.04); NSW of Department of Education (2018247); and the Maitland-Newcastle, Sydney, Wollongong, Bathurst, Parramatta, Wagga Wagga and Canberra-Goulburn Catholic Dioceses. The trial is prospectively registered with the Australian New Zealand Clinical Trials Registry (ACTRN12623000558628). Consent from schools to participate in the data collection component of the study (i.e. the follow-up survey) will be obtained with approved information statements and consent processes. Participants may withdraw from the study at any time. Evaluation and process data collected as part of the study will be disseminated widely through national and international peer-reviewed publications and conferences presentations.

### Data management

The participant information statement informs participants about the planned or possible future use of information/data. All hard copy information will be stored at the workplace of the research team at Hunter New England Population Health’s secure Wallsend location in locked filing cabinets and secure computer files. Only research personnel and approved staff working with the data will have access to the data. Any electronic data will only be accessible via password protected accounts and any file-sharing will be restricted to members of the project team. All identifying information will be kept for the required 7 years until it needs to be destroyed.

In the interest of open science we would like to be able to share de-identified data and provide access to any potential systematic reviews at the individual level as well as any required secondary analysis. Other researchers may seek access to the data for the purposes of re-analysis and secondary analysis. Any use of data that is not covered by the current ethics approval will require additional ethics approval before the data is made available.

Anyone seeking to access the data will need to contact the lead investigator, along with seeking appropriate ethical clearances. Only once those approvals are granted will de-identified data be shared via an encrypted communication channel.

### Consent for publication

Not applicable.

## SUPPLEMENTARY FILE 1

**SPIRIT 2013 Checklist: Recommended items to address in a clinical trial protocol and related documents**

SPRIT 2013 Checklist: Recommended items to address in a clinical trial protocol and related documents
| Section/item | Item No | Description | Addressed on page number |
| --- | --- | --- | --- |
| **Administrative information** |  |  |  |
| Title | 1 | Descriptive title identifying the study design, population, interventions, and, if applicable, trial acronym | 1 |
| Trial registration | 2a | Trial identifier and registry name. If not yet registered, name of intended registry | 8 |
|  | 2b | All items from the World Health Organization Trial Registration Data Set | 8 |
| Protocol version | 3 | Date and version identifier | 1 |
| Funding | 4 | Sources and types of financial, material, and other support | 18 |
| Roles and responsibilities | 5a | Names, affiliations, and roles of protocol contributors | 1-2 |
|  | 5b | Name and contact information for the trial sponsor | 18 |
|  | 5c | Role of study sponsor and funders, if any, in study design; collection, management, analysis, and interpretation of data; writing of the report; and the decision to submit the report for publication, including whether they will have ultimate authority over any of these activities | 18 |
|  | 5d | Composition, roles, and responsibilities of the coordinating centre, steering committee, endpoint adjudication committee, data management team, and other individuals or groups overseeing the trial, if applicable (see Item 21a for data monitoring committee) | 17 |
| **Introduction** |  |  |  |
| Background and rationale | 6a | Description of research question and justification for undertaking the trial, including summary of relevant studies (published and unpublished) examining benefits and harms for each intervention | 5-7 |
|  | 6b | Explanation for choice of comparators | 9-12 |
| Objectives | 7 | Specific objectives or hypotheses | 7 |
| Trial design | 8 | Description of trial design including type of trial (eg, parallel group, crossover, factorial, single group), allocation ratio, and framework (eg, superiority, equivalence, noninferiority, exploratory) | 7 |
| **Methods: Participants, interventions, and outcomes** |  |  |  |
| Study setting | 9 | Description of study settings (eg, community clinic, academic hospital) and list of countries where data will be collected. Reference to where list of study sites can be obtained | 8 |
| Eligibility criteria | 10 | Inclusion and exclusion criteria for participants. If applicable, eligibility criteria for study centres and individuals who will perform the interventions (eg, surgeons, psychotherapists) | 9 |
| Interventions | 11a | Interventions for each group with sufficient detail to allow replication, including how and when they will be administered | 9-12 |
|  | 11b | Criteria for discontinuing or modifying allocated interventions for a given trial participant (eg, drug dose change in response to harms, participant request, or improving/worsening disease) | N/A |
|  | 11c | Strategies to improve adherence to intervention protocols, and any procedures for monitoring adherence (eg, drug tablet return, laboratory tests) | 12 |
|  | 11d | Relevant concomitant care and interventions that are permitted or prohibited during the trial | N/A |
| Outcomes | 12 | Primary, secondary, and other outcomes, including the specific measurement variable (eg, systolic blood pressure), analysis metric (eg, change from baseline, final value, time to event), method of aggregation (eg, median, proportion), and time point for each outcome. Explanation of the clinical relevance of chosen efficacy and harm outcomes is strongly recommended | 15-16 |
| Participant timeline | 13 | Time schedule of enrolment, interventions (including any run-ins and washouts), assessments, and visits for participants. A schematic diagram is highly recommended (see Figure) | N/A |
| Sample size | 14 | Estimated number of participants needed to achieve study objectives and how it was determined, including clinical and statistical assumptions supporting any sample size calculations | 16 |
| Recruitment | 15 | Strategies for achieving adequate participant enrolment to reach target sample size | 8-9 |
| **Methods: Assignment of interventions (for controlled trials)** |  |  |  |
| Allocation: |  |  |  |
| Sequence generation | 16a | Method of generating the allocation sequence (eg, computer-generated random numbers), and list of any factors for stratification. To reduce predictability of a random sequence, details of any planned restriction (eg, blocking) should be provided in a separate document that is unavailable to those who enrol participants or assign interventions | 9-10 |
| Allocation concealment mechanism | 16b | Mechanism of implementing the allocation sequence (eg, central telephone; sequentially numbered, opaque, sealed envelopes), describing any steps to conceal the sequence until interventions are assigned | 9-10 |
| Implementation | 16c | Who will generate the allocation sequence, who will enrol participants, and who will assign participants to interventions | 9-10 |
| Blinding (masking) | 17a | Who will be blinded after assignment to interventions (eg, trial participants, care providers, outcome assessors, data analysts), and how | 9-10 |
|  | 17b | If blinded, circumstances under which unblinding is permissible, and procedure for revealing a participant's allocated intervention during the trial | 9-10 |
| **Methods: Data collection, management, and analysis** |  |  |  |
| Data collection methods | 18a | Plans for assessment and collection of outcome, baseline, and other trial data, including any related processes to promote data quality (eg, duplicate measurements, training of assessors) and a description of study instruments (eg, questionnaires, laboratory tests) along with their reliability and validity, if known. Reference to where data collection forms can be found, if not in the protocol | 15-16 |
|  | 18b | Plans to promote participant retention and complete follow-up, including list of any outcome data to be collected for participants who discontinue or deviate from intervention protocols | 15-16 |
| Data management | 19 | Plans for data entry, coding, security, and storage, including any related processes to promote data quality (eg, double data entry; range checks for data values). Reference to where details of data management procedures can be found, if not in the protocol | 19 |
| Statistical methods | 20a | Statistical methods for analysing primary and secondary outcomes. Reference to where other details of the statistical analysis plan can be found, if not in the protocol | 16-17 |
|  | 20b | Methods for any additional analyses (eg, subgroup and adjusted analyses) | 16-17 |
|  | 20c | Definition of analysis population relating to protocol non-adherence (eg, as randomised analysis), and any statistical methods to handle missing data (eg, multiple imputation) | 16-17 |
| **Methods: Monitoring** |  |  |  |
| Data monitoring | 21a | Composition of data monitoring committee (DMC); summary of its role and reporting structure; statement of whether it is independent from the sponsor and competing interests; and reference to where further details about its charter can be found, if not in the protocol. Alternatively, an explanation of why a DMC is not needed | 17 |
|  | 21b | Description of any interim analyses and stopping guidelines, including who will have access to these interim results and make the final decision to terminate the trial | N/A |
| Harms | 22 | Plans for collecting, assessing, reporting, and managing solicited and spontaneously reported adverse events and other unintended effects of trial interventions or trial conduct | N/A |
| Auditing | 23 | Frequency and procedures for auditing trial conduct, if any, and whether the process will be independent from investigators and the sponsor | N/A |
| **Ethics and dissemination** |  |  |  |
| Research ethics approval | 24 | Plans for seeking research ethics committee/institutional review board (REC/IRB) approval | 8 |
| Protocol amendments | 25 | Plans for communicating important protocol modifications (eg, changes to eligibility criteria, outcomes, analyses) to relevant parties (eg, investigators, REC/IRBs, trial participants, trial registries, journals, regulators) | N/A |
| Consent or assent | 26a | Who will obtain informed consent or assent from potential trial participants or authorised surrogates, and how (see Item 32) | 9 |
|  | 26b | Additional consent provisions for collection and use of participant data and biological specimens in ancillary studies, if applicable | N/A |
| Confidentiality | 27 | How personal information about potential and enrolled participants will be collected, shared, and maintained in order to protect confidentiality before, during, and after the trial | 19 |
| Declaration of interests | 28 | Financial and other competing interests for principal investigators for the overall trial and each study site | 18 |
| Access to data | 29 | Statement of who will have access to the final trial dataset, and disclosure of contractual agreements that limit such access for investigators | 19 |
| Ancillary and post-trial care | 30 | Provisions, if any, for ancillary and post-trial care, and for compensation to those who suffer harm from trial participation | N/A |
| Dissemination policy | 31a | Plans for investigators and sponsor to communicate trial results to participants, healthcare professionals, the public, and other relevant groups (eg, via publication, reporting in results databases, or other data sharing arrangements), including any publication restrictions | 18-19 |
|  | 31b | Authorship eligibility guidelines and any intended use of professional writers | Supplementary materials |
|  | 31c | Plans, if any, for granting public access to the full protocol, participant-level dataset, and statistical code | 19 |
| **Appendices** |  |  |  |
| Informed consent materials | 32 | Model consent form and other related documentation given to participants and authorised surrogates | Supplementary materials |
| Biological specimens | 33 | Plans for collection, laboratory evaluation, and storage of biological specimens for genetic or molecular analysis in the current trial and for future use in ancillary studies, if applicable | N/A |

## Notes

### Competing Interest Statement

The authors have declared no competing interest.

### Clinical Trial

Australian New Zealand Clinical Trials Registry (ANZCTR): 12623000558628

## REFERENCES

1. Australian Institute of Health and Welfare. Burden of disease. Canberra: Australian Government; 2020.

2. Steinberger J, Daniels SR, Hagberg N, Isasi CR, Kelly AS, Lloyd-Jones D, et al. Cardiovascular Health Promotion in Children: Challenges and Opportunities for 2020 and Beyond: A Scientific Statement From the American Heart Association. Circulation. 2016;134(12):e236–e55.

3. Australian Institute of Health and Welfare. Nutrition across the life stages. Canberra: AIHW; 2018.

4. Angell SY, McConnell MV, Anderson CAM, Bibbins-Domingo K, Boyle DS, Capewell S, et al. The American Heart Association 2030 Impact Goal: A Presidential Advisory From the American Heart Association. Circulation. 2020;141(9):e120–e38.

5. United Nations. Decade Action on Nutrition 2023 [cited 2023 16 October]. Available from: https://www.un.org/nutrition/.

6. Moodie AR. Australia: the healthiest country by 2020. Medical Journal of Australia. 2008;189(10):588-90.

7. Sanigorski AM, Bell AC, Kremer PJ, Swinburn BA. Lunchbox contents of Australian school children: room for improvement. European journal of clinical nutrition. 2005;59(11):1310–6.

8. Reynolds R, Sutherland R, Nathan N, Janssen L, Lecathelinais C, Reilly K, et al. Feasibility and principal acceptability of school-based mobile communication applications to disseminate healthy lunchbox messages to parents. Health Promotion Journal of Australia. 2019;30(1):108–13.

9. Nathan N, Janssen L, Sutherland R, Hodder RK, Evans CEL, Booth D, et al. The effectiveness of lunchbox interventions on improving the foods and beverages packed and consumed by children at centre-based care or school: a systematic review and meta-analysis. The international journal of behavioral nutrition and physical activity. 2019;16(1):38.

10. Barnes C, Hall A, Nathan N, Sutherland R, McCarthy N, Pettet M, et al. Efficacy of a school-based physical activity and nutrition intervention on child weight status: Findings from a cluster randomized controlled trial. Prev Med. 2021;153:106822.

11. Sutherland R, Nathan N, Brown A, Yoong S, Finch M, Lecathelinais C, et al. A randomized controlled trial to assess the potential efficacy, feasibility and acceptability of an m-health intervention targeting parents of school aged children to improve the nutritional quality of foods packed in the lunchbox ’SWAP IT’. The international journal of behavioral nutrition and physical activity. 2019;16(1):54.

12. National Health and Medical Research Council. Australian Dietary Guidelines. Canberra: National Health and Medical Research Council; 2013.

13. Wolfenden L, McCrabb S, Barnes C, O’Brien KM, Ng KW, Nathan NK, et al. Strategies for enhancing the implementation of school-based policies or practices targeting diet, physical activity, obesity, tobacco or alcohol use. Cochrane Database of Systematic Reviews. 2022(8).

14. Diderichsen F. The relevance of public health research for practice: A 30-year perspective. Scandinavian journal of public health. 2018;46(22_suppl):58-66.

15. Oh A, Abazeed A, Chambers DA. Policy Implementation Science to Advance Population Health: The Potential for Learning Health Policy Systems. Frontiers in public health. 2021;9:681602.

16. Woodcock J, LaVange LM. Master Protocols to Study Multiple Therapies, Multiple Diseases, or Both. New England Journal of Medicine. 2017;377(1):62–70.

17. Meyer EL, Mesenbrink P, Dunger-Baldauf C, Fülle HJ, Glimm E, Li Y, et al. The Evolution of Master Protocol Clinical Trial Designs: A Systematic Literature Review. Clinical therapeutics. 2020;42(7):1330–60.

18. Park JJH, Siden E, Zoratti MJ, Dron L, Harari O, Singer J, et al. Systematic review of basket trials, umbrella trials, and platform trials: a landscape analysis of master protocols. Trials. 2019;20(1):572.

19. Seidler AL, Hunter KE, Cheyne S, Ghersi D, Berlin JA, Askie L. A guide to prospective meta-analysis. BMJ. 2019;367:l5342.

20. National Centre of Implementation Science. About NCOIS 2023 [cited 2023 12 October]. Available from: https://ncois.org.au/.

21. Barnes C, Sutherland R, Jones G, Kingon N, Collaborative NR, Wolfenden L. Development and piloting of a Community of Practice to support learning and improvement in health promotion practice within NSW local health districts. Public Health Research & Practice. 2023.

22. Innes-Hughes C. BA, Buffett K., Henderson L., Lockeridge A., Pimenta N., Radvan D., Rissel C. NSW Healthy Children Initiative: The first five years July 2011 – June 2016. NSW Ministry of Health; 2017.

23. National Centre of Implementation Science. GREAT-HP 2023 Stakeholder Survey Preliminary Findings. Newcastle: University of Newcastle; 2023.

24. Cuschieri S. The CONSORT statement. Saudi journal of anaesthesia. 2019;13(Suppl 1):S27–s30.

25. Chan A-W, Tetzlaff JM, Gøtzsche PC, Altman DG, Mann H, Berlin JA, et al. SPIRIT 2013 explanation and elaboration: guidance for protocols of clinical trials. BMJ : British Medical Journal. 2013;346:e7586.

26. Australian Bureau of Statistics (ABS). Statistical Geography Volume 1 - Australian Geographical Classification (ASGC), . Canberra: Commonwealth of Australia; 2022.

27. Australian Bureau of Statistics (ABS). Socio-Economic Indexes for Areas (SEIFA) Australia 2021 [cited 2023 16 October]. Available from: https://www.abs.gov.au/statistics/people/people-and-communities/socio-economic-indexes-areas-seifa-australia/latest-release#:~:text=SEIFA%20combines%20Census%20data%20such,is%20compared%20with%20other%20areas.

28. Yoong SL, Nathan NK, Wyse RJ, Preece SJ, Williams CM, Sutherland RL, et al. Assessment of the School Nutrition Environment: A Study in Australian Primary School Canteens. American Journal of Preventive Medicine. 2015;49(2):215–22.

29. Barnes C, Sutherland R, Jones J, Brown A, Stacey F, Wolfenden L. Maximising the adoption of a school-based m-Health intervention to improve the nutritional quality of student lunchboxes to ensure population-level impact. Health promotion journal of Australia : official journal of Australian Association of Health Promotion Professionals. 2022;33 Suppl 1:412–4.

30. Wisdom JP, Chor KH, Hoagwood KE, Horwitz SM. Innovation adoption: a review of theories and constructs. Administration and policy in mental health. 2014;41(4):480–502.

31. Powell BJ, Waltz TJ, Chinman MJ, Damschroder LJ, Smith JL, Matthieu MM, et al. A refined compilation of implementation strategies: results from the Expert Recommendations for Implementing Change (ERIC) project. Implementation Science. 2015;10(1):21.

32. Michie S, Richardson M, Johnston M, Abraham C, Francis J, Hardeman W, et al. The behavior change technique taxonomy (v1) of 93 hierarchically clustered techniques: building an international consensus for the reporting of behavior change interventions. Annals of behavioral medicine : a publication of the Society of Behavioral Medicine. 2013;46(1):81–95.

33. Weiner BJ, Lewis CC, Stanick C, Powell BJ, Dorsey CN, Clary AS, et al. Psychometric assessment of three newly developed implementation outcome measures. Implementation Science. 2017;12(1):108.

34. Australian Curriculum Assessment and Reporting Authority. MySchool 2023 [cited 2023 12 October]. Available from: https://www.myschool.edu.au/.

35. Riley RD, Dias S, Donegan S, Tierney JF, Stewart LA, Efthimiou O, et al. Using individual participant data to improve network meta-analysis projects. BMJ evidence-based medicine. 2023;28(3):197–203.

